# Automatic Retinoblastoma Screening and Surveillance Using Deep Learning

**DOI:** 10.1101/2022.08.23.22279103

**Authors:** Ruiheng Zhang, Li Dong, Ruyue Li, Kai Zhang, Yitong Li, Hongshu Zhao, Jitong Shi, Xin Ge, Xiaolin Xu, Libin Jiang, Xuhan Shi, Chuan Zhang, Wenda Zhou, Liangyuan Xu, Haotian Wu, Heyan Li, Chuyao Yu, Jing Li, Jianmin Ma, Wenbin Wei

## Abstract

Retinoblastoma is the most common intraocular malignancy in childhood. With the advanced management strategy, the global salvage and overall survival have significantly improved, which proposes subsequent challenges regarding long-term surveillance and offspring screening. Here, we developed deep learning algorithm, called Deep Learning Assistant for Retinoblastoma (DLA-RB) training on A total of 36623 images from 713 patients. We validated it in the prospectively collected dataset, comprised of 1366 images form 139 eyes of 103 patients. In identifying active retinoblastoma tumors (treatment required) from all clinical-suspected patients, the area under the receiver operating characteristic curve (AUC) of DLA-RB reached 0.991 (95% CI 0.970-1.000). In identifying active retinoblastoma from stable retinoblastoma patients (treatment is not required), AUC of DLA-RB reached 0.962 (95% CI 0.915-1.000), respectively. Cost-utility analysis revealed that DLA-RB based diagnosis mode is more cost-effective in both retinoblastoma diagnosis and retinoblastoma activity surveillance. The DLA-RB achieved high accuracy and sensitivity in identifying active retinoblastoma from the normal and stable retinoblastoma fundus. It can be incorporated within telemedicine programs in the future.

## Introduction

Retinoblastoma is the most common intraocular malignancy in childhood. It is estimated to affect one case per 16,000–18,000 live births worldwide.^1^ China, India, and other populous countries in Asia and Africa have the largest amount of newly diagnosed retinoblastoma patients. With advanced screening techniques and multidisciplinary management, most retinoblastoma patients achieve life-saving, globe salvage, and even useful vision.^2^ In a recent national cohort study in China, the overall survival rates of RB patients were 81%, 83%, and 91% in patients diagnosed in 1989-2008, 2009-2013, 2014-2017, respectively.^3^ For globe salvage, almost all retinoblastoma classified by the International Intraocular Retinoblastoma Classification (IIRC)^4^ as group A-C can achieve globe salvage through systemic chemotherapy with adjuvant laser therapy and cryotherapy.^5^ Group D retinoblastoma can also achieve 40% (95%CI:31–51%) globe salvage rate through chemotherapy alone ^6^ and a significantly higher globe salvage rate when receiving intra-ophthalmic artery chemotherapy.^7^

Improved overall survival and successful local control propose subsequent challenges regarding long-term surveillance after tumor control and screening for the offspring and relatives of retinoblastoma survivors. The current expert consensus recommends every 2–6 months for 4 years after tumor control.^1^ In real-world practice, group D retinoblastoma received an average of 21 examinations under anesthesia (EUAs) during 5-year follow-up.^8^ Furthermore, offspring of retinoblastoma survivors have 50% chance of inheriting the mutant RB1 from the affected parent, which would result in a 97% risk of developing retinoblastoma.^9^ Global preserving approach thus leads to additional disease burden comprised of health care, socioeconomic, and mental aspects. Furthermore, in most developing countries, only experienced medical ophthalmologists at tertiary eye centers can perform EUAs. This unbalanced distribution of health care resources may delay the diagnosis and proper management during the referring process.

The application of deep learning (DL) techniques proposes reliable and low-cost methods for screening and diagnosing retinal diseases. Previous studies showed that the DL algorithm based on fundus images achieves close-to-expert performance for the automatic detection of retinal diseases, including diabetic retinopathy^10^, age-related macular degeneration (AMD),^11^ glaucoma,^12^ myopic maculopathy,^13^ rand papilledema.^14^ Recently, researchers have applied DL in detecting plus diseases in retinopathy of prematurity.^15,16^ The DL algorithm exceeds 6 of 8 experts in identifying plus diseases, which indicates it can reach robust performance in images obtained by EUAs.

Thus, in the current study, we developed Deep Learning Assistant for Retinoblastoma (DLA-RB) and validated it in the prospectively collected dataset. The DLA-RB aims to assist the fundus surveillance after local control and provide referral advice. The DLA-RB also provides automatic surveillance of the contralateral eye of retinoblastoma patients and the offspring of retinoblastoma survivors.

## Results

Between March 2018 and January 2022, 47503 images from 713 patients were retrospectively collected from the anonymous data center in Beijing Tongren hospital (**Figure 1**). After manually excluding low-quality images due to non-fundus image, halation, blurs, and defocus (**Supplementary Figure S1**), a total of 36623 images from 713 patients were finally included for DLA-RB development, with 19045 (52.0%), 2918 (8.0%), and 14660 (40.0%) images were normal, stable and active retinoblastoma (**Supplementary Table S1**). At patient levels, 713 patients were randomly allocated for DLA-RB development and five-fold cross-validation. Considering that it is more complicated to distinguish between stable and active retinoblastoma, we first trained ResNet-50 and InceptionResnetV3 for this two-class task to compare the performance of these two architectures. ROC indicated slight better performance of ResNet-50 in five-fold cross-validation (0.940 [95%CI 0.851-0.996] vs. 0.934 [95%CI 0.735-0.946], **Supplementary Figure S2**). Thus, ResNet-50 was used to establish DLA-RB. To distinguish between the normal fundus and active lesion, DLA-RB achieved an AUC of 0.9982 (95%CI 0.986-1.000) in the development dataset (**Supplementary Figure S3**).

**Figure 1.**
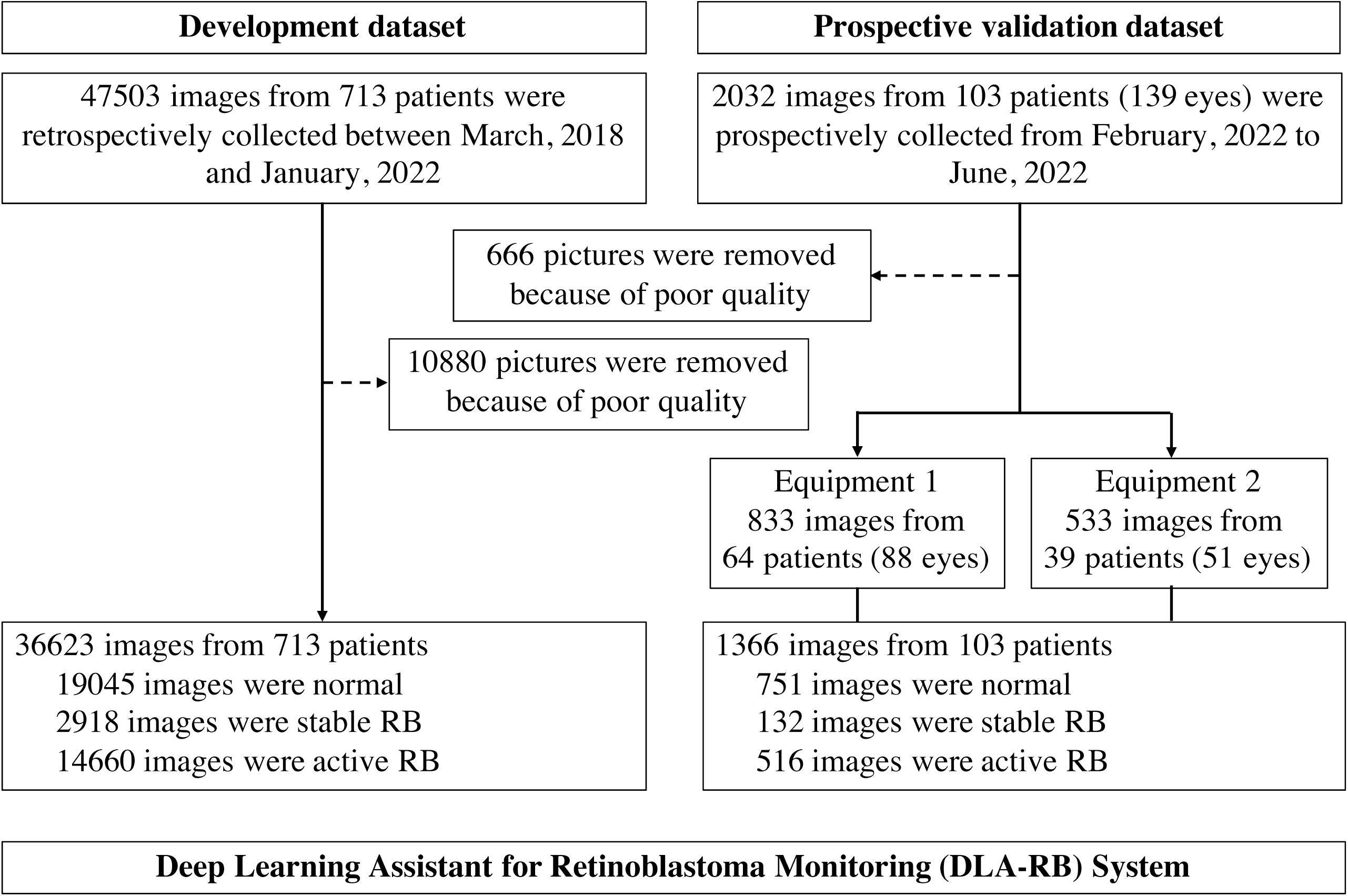
Workflow diagram for the development and evaluation of DLA-RB. RB, retinoblastoma.

From February 2022 to June 2022, 103 patients with clinical-suspected retinoblastoma and treated retinoblastoma patients first visited Beijing Tongren hospitals. Fundus images of these patients were not used in developing DLA-RB. If more than one time of EUA, only the first EUA images were collected. In total, 139 eyes of 103 patients were included for prospective validation (**Table 1**). 69 eyes were clinically diagnosed with retinoblastoma. According to international intraocular retinoblastoma classification (IIRC),^17^ 3 (4.3%), 7 (10.1%), 8 (11.6%), 40 (58.0%), and 11 (15.9%) eye was classified as A-E stage, respectively. 43.5% of retinoblastoma was endophytic, while 56.5% were exophytic. For each EUA, the highest probability among all images was assigned as an eye-level probability.

**Table 1.** Basic characteristics of patients in prospective validation dataset.

|  | Equipment 1 | Equipment 2 |
| --- | --- | --- |
| Number of patients | 64 | 39 |
| Age at diagnosis |  |  |
| <12month | 14 (21.9%) | 11 (28.2%) |
| 12-23 month | 19 (29.7%) | 15 (38.5%) |
| 24-35 month | 14 (21.9%) | 6 (15,4%) |
| ≥36 month | 17 (26.6%) | 7 (17.9%) |
| Gender |  |  |
| male | 36 (56.3%) | 23 (59.0%) |
| female | 28 (43.8%) | 16 (41.0%) |
| Laterality |  |  |
| unilateral | 44 (68.7%) | 22 (56.4%) |
| bilateral | 19 (29.7%) | 16 (41.0%) |
| Number of eyes | 88 | 51 |
| IIRC Grade |  |  |
| normal fundus | 46 (52.3%) | 24 (47.1%) |
| A | 3 (3.4%) | 0 (0.0%) |
| B | 5 (5.7%) | 2 (3.9%) |
| C | 6 (6.8%) | 2 (3.9%) |
| D | 22 (25.0%) | 18 (35.3%) |
| E | 6 (6.8%) | 5 (9.8%) |
| Number of eyes with retinoblastoma | 42 | 27 |
| Tumor focality |  |  |
| unifocal | 16 (38.1%) | 4 (14.8%) |
| multifocal | 26 (61.9%) | 23 (85.2%) |
| Retinoblastoma growth pattern |  |  |
| endophytic | 24 (57.1%) | 15 (55.6%) |
| exophytic | 18 (42.9%) | 12 (44.4%) |
| Fovea involvement |  |  |
| not involved | 14 (33.3%) | 6 (22.2%) |
| subfoveal fluid | 2 (4.8%) | 4 (14.8%) |
| foveal tumor | 26 (61.9%) | 17 (63.0%) |
| Retinoblastoma seeds |  |  |
| not seeds | 17 (40.5%) | 7 (25.9%) |
| subretinal | 16 (38.1%) | 10 (37.0%) |
| vitreous | 9 (21.4%) | 10 (37.0%) |

IIRC, international intraocular retinoblastoma classification.

DLA-RB could accurately identify active retinoblastoma tumors from all clinical-suspected retinoblastoma (**Figure 2A-B, Table 2**). The AUC, sensitivity, and specificity reached 0.991 (95%CI 0.970-1.000), 0.979 (95%CI 0.927-1.000), and 1.000 (95%CI 1.000-1.000), respectively. Compared with the human ophthalmologists, the DLA-RB reached significantly higher sensitivity and specificity in identifying new retinoblastoma tumors (**Figure 2A-B, Supplementary Table S2-3)**. The only misclassified case of DLA-RB was a Group A retinoblastoma located at superior-nasal ora Serrata (**Supplementary Figure S1)**.

**Table 2.**
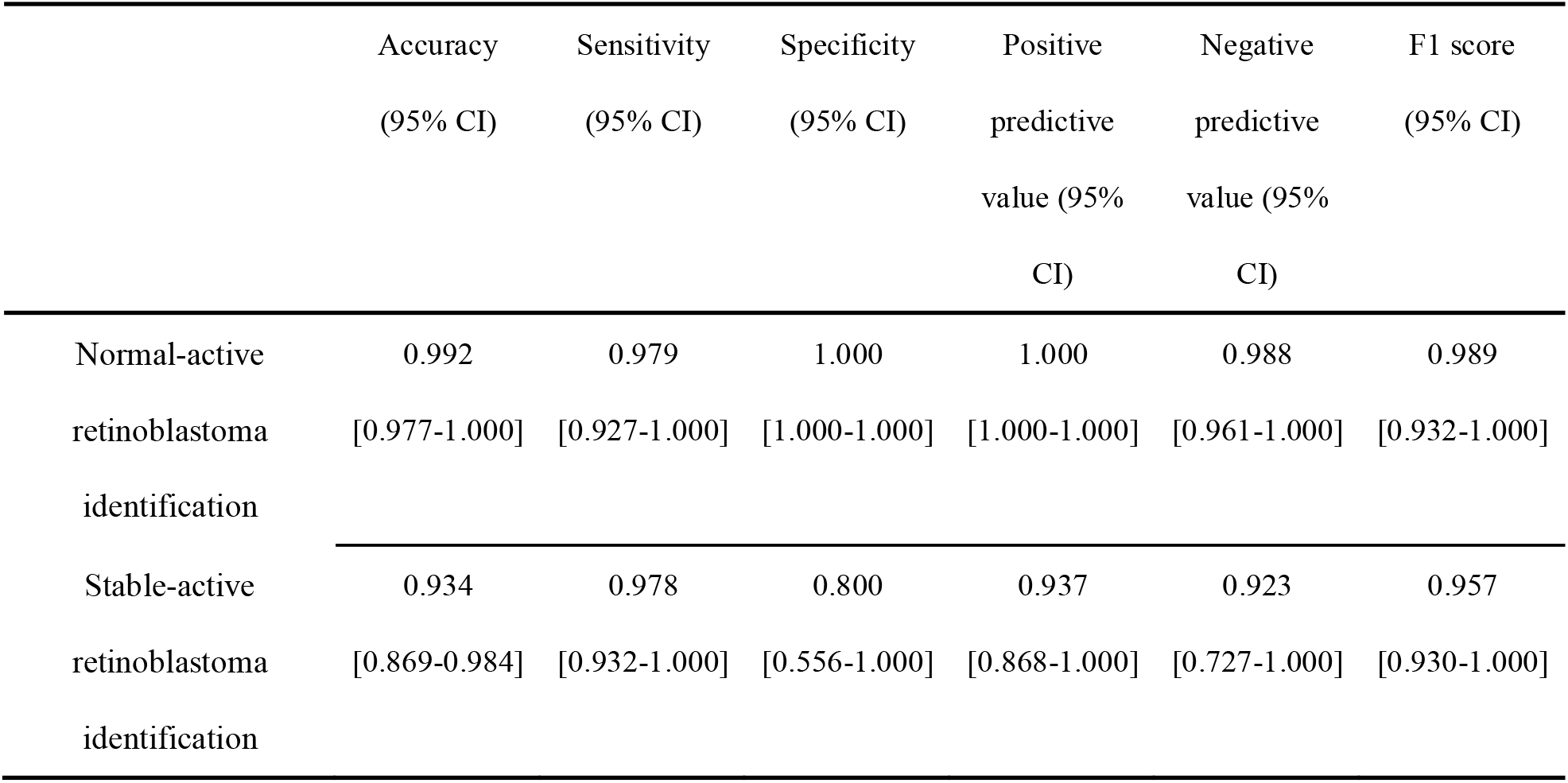
Performance of DLA-RB in prospective validation dataset,

|  | Accuracy<br>(95% CI) | Sensitivity<br>(95% CI) | Specificity<br>(95% CI) | Positive<br>predictive<br>value (95%<br>CI) | Negative<br>predictive<br>value (95%<br>CI) | F1 score<br>(95% CI) |
| --- | --- | --- | --- | --- | --- | --- |
| Normal-active<br>retinoblastoma<br>identification | 0.992<br>[0.977-1.000] | 0.979<br>[0.927-1.000] | 1.000<br>[1.000-1.000] | 1.000<br>[1.000-1.000] | 0.988<br>[0.961-1.000] | 0.989<br>[0.932-1.000] |
| Stable-active<br>retinoblastoma<br>identification | 0.934<br>[0.869-0.984] | 0.978<br>[0.932-1.000] | 0.800<br>[0.556-1.000] | 0.937<br>[0.868-1.000] | 0.923<br>[0.727-1.000] | 0.957<br>[0.930-1.000] |

**Figure 2.**
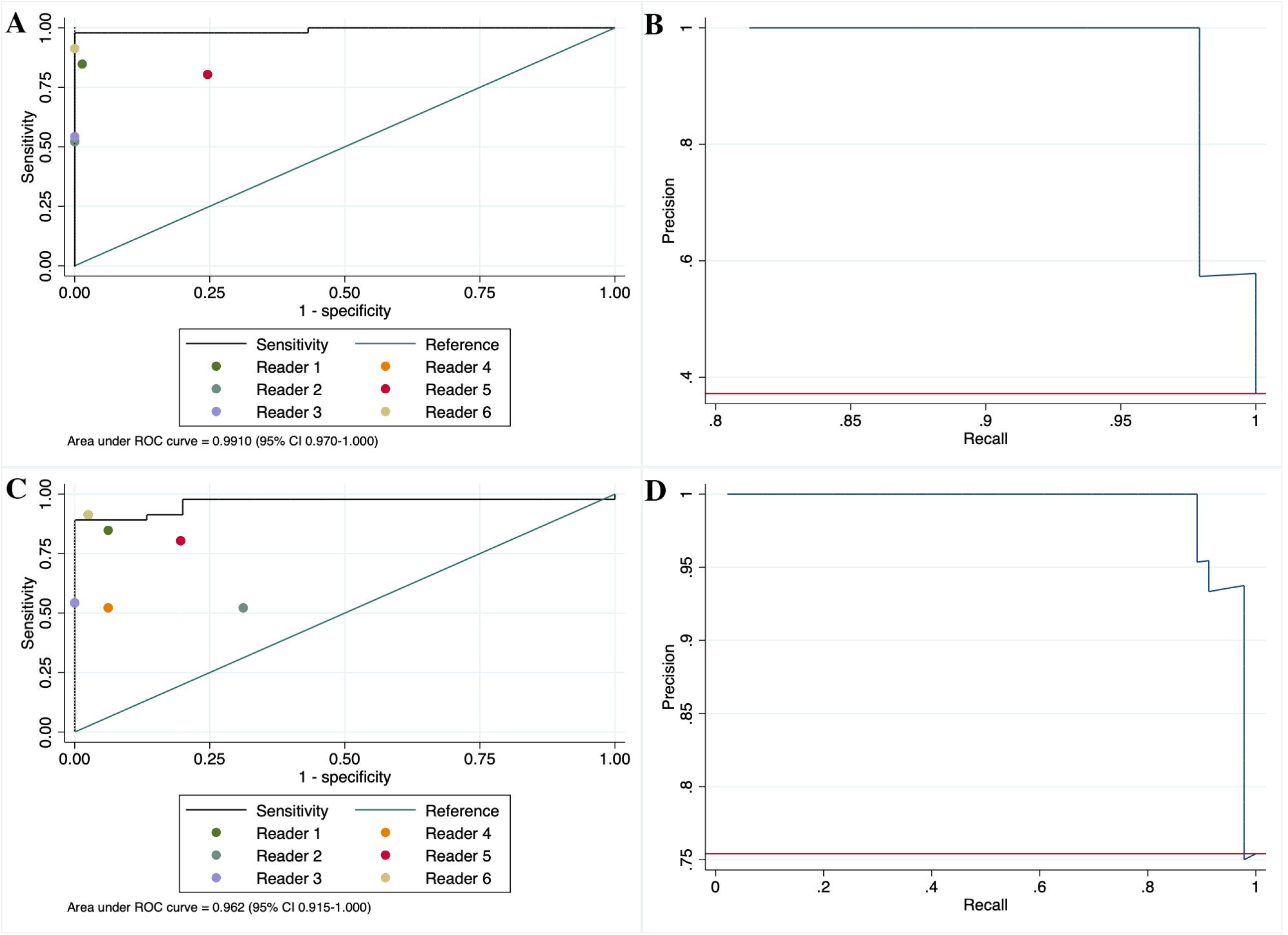
Receiver operating characteristic curves and precision-recall curve of DLA-RB performance in prospective validation dataset. Receiver operating characteristic and precision-recall curve of DLA-RB in identifying normal-active lesion identification (A-B), and stable-active lesion identification (C-D).

DLA-RB could also accurately distinguish active retinoblastoma from stable retinoblastoma (**Figure 2C-D, Table 2**). Among all referred and treated retinoblastoma patients, the AUC, sensitivity, and specificity reached 0.962 (95%CI 0.915-1.000), 0.978 (95%CI 0.932-1.000), and 0.800 (95%CI 0.556-1.000) respectively. Compared with competent ophthalmologists, the DLA-RB reached superior sensitivity yet inferior specificity in identifying active retinoblastoma from stable retinoblastoma (**Figure 2C-D, Supplementary Table S2-3)**. Of all 4 misclassified cases in this task, 3 cases were false-positive, whereas only 1 case was false-negative (**Table 4)**.

**Table 3.** Confusion matrix of DLA-RB in identifying active retinoblastoma among treated patients.

|  |  | DLA-RB |  |  |
| --- | --- | --- | --- | --- |
|  |  | stable | active | total |
| Gold<br>standard | stable | 12 | 3 | 15 |
|  | active | 1 | 45 | 46 |
|  | total | 13 | 48 | 61 |

**Table 4.**
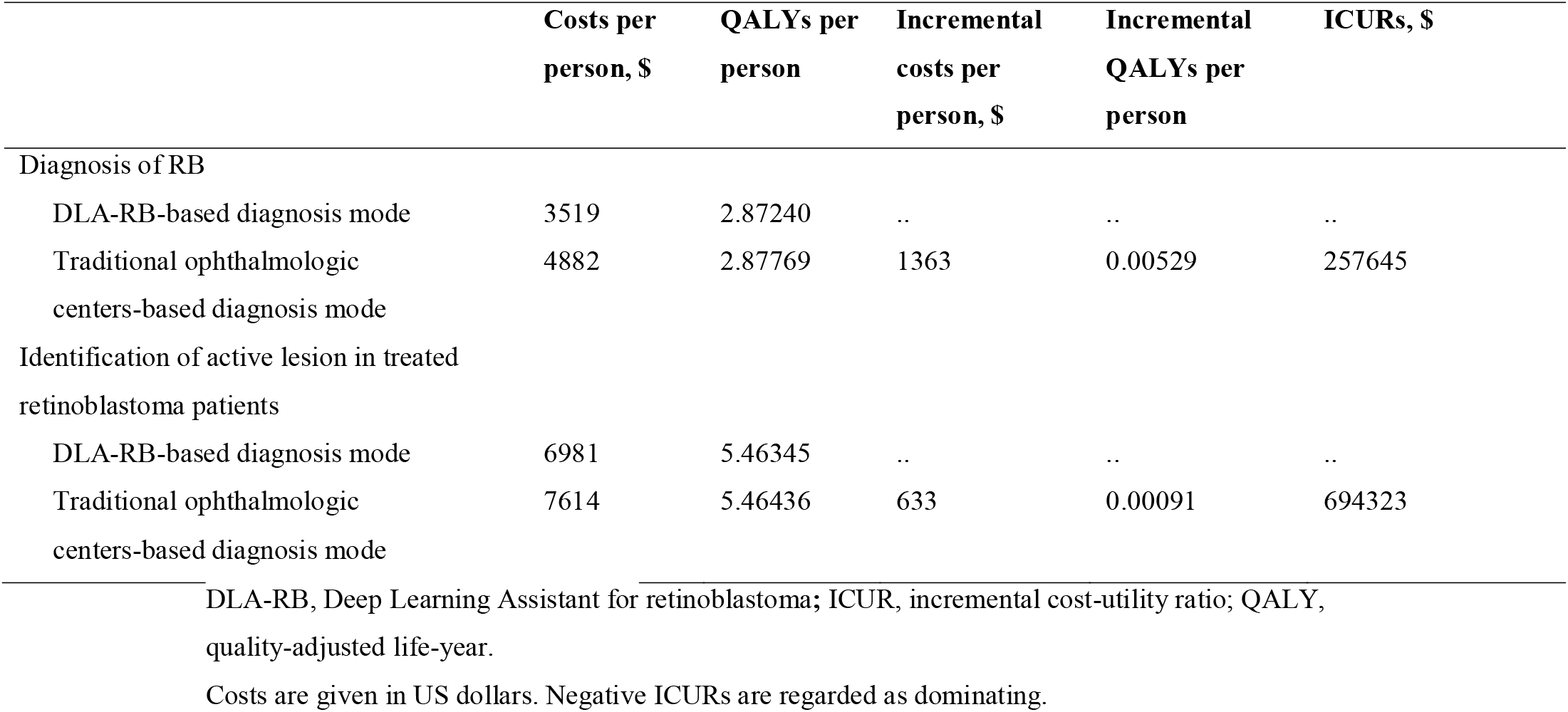
Base-case cost-utility results.

| | Costs per<br>person, \$ | QALYs per<br>person | Incremental<br>costs per<br>person, \$ | Incremental<br>QALYs per<br>person | ICURs, \$ |
| --- | --- | --- | --- | --- | --- |
| Diagnosis of RB |  |  |  |  |  |
| DLA-RB-based diagnosis mode | 3519 | 2.87240 | .. | .. | .. |
| Traditional ophthalmologic<br>centers-based diagnosis mode | 4882 | 2.87769 | 1363 | 0.00529 | 257645 |
| Identification of active lesion in treated<br>retinoblastoma patients |  |  |  |  |  |
| DLA-RB-based diagnosis mode | 6981 | 5.46345 | .. | .. | .. |
| Traditional ophthalmologic<br>centers-based diagnosis mode | 7614 | 5.46436 | 633 | 0.00091 | 694323 |

DLA-RB, Deep Learning Assistant for retinoblastoma; ICUR, incremental cost-utility ratio; QALY,
quality-adjusted life-year.
Costs are given in US dollars. Negative ICURs are regarded as dominating.

Heatmap visualization revealed that the DLA-RB mainly focused on the tumor of the newly-diagnosed retinoblastoma (**Figure 3A**). The DLA-RB focused on the relapse site of retinoblastoma on fundus images (**Figure 3B)**.

**Figure 3.**
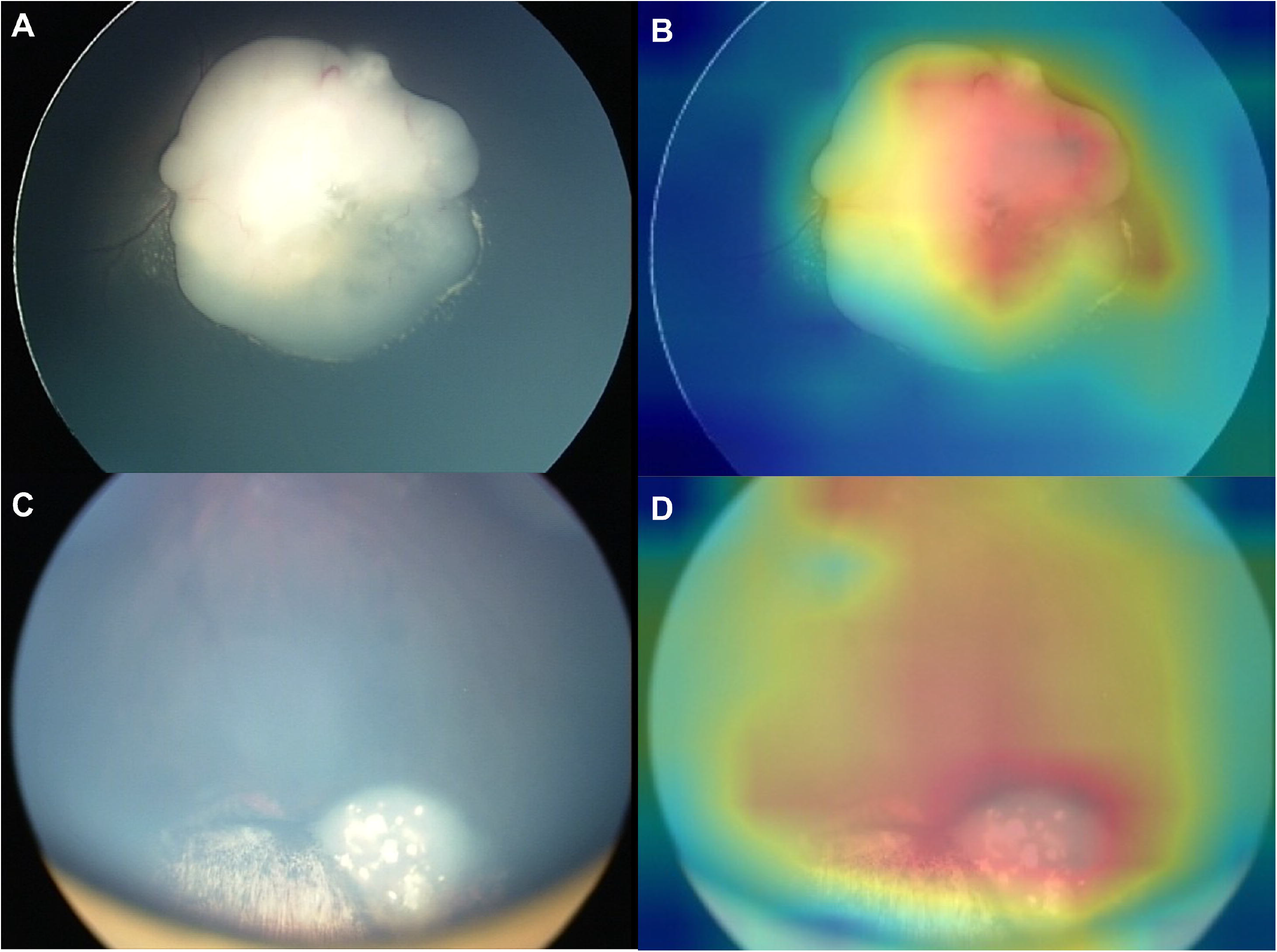
Heatmap Visualization of DLA-RB Heatmap demonstrating representative lesions, shown in original fundus image (first column), general heatmap (second column) for (A) identifying active retinoblastoma tumors from all clinical-suspected retinoblastoma and (B) identifying active retinoblastoma from all referred and treated retinoblastoma patients.

DLA-DR has proven to be sensitive and accuracy in identifying active retinoblastoma from the normal fundus and stable retinoblastoma fundus. Yet, the cost-effectiveness of a DLA-RB based approach to screen and surveillance retinoblastoma is unknown. **Table 4** shows the base-case results of the cost-utility analysis. For identifying active retinoblastoma tumors from all clinical-suspected retinoblastoma, the cumulative costs for traditional ophthalmologic centers-based diagnosis mode and DLA-RB based diagnosis mode were $4882 and $3519, respectively, and cumulative quality-adjusted life-year (QALYs) for both diagnosis modes were 2.87769 and 2.8724 respectively. Compared with DLA-RB based diagnosis mode, the traditional ophthalmologic centers-based diagnosis mode was not cost-effective since it resulted in a gain of 1 QALY at the cost of $257645. For distinguishing active retinoblastoma from stable retinoblastoma patients, the cumulative costs and QALYs for traditional ophthalmologic centers-based diagnosis mode were $7614 and 5.46436, and the figures for DLA-RB-based diagnosis mode were $6981 and 5.46345. Traditional ophthalmologic centers-based diagnosis mode was still unsatisfied with the cost-effectiveness threshold since it produced an ICUR of $694323. Therefore, DLA-RB based diagnosis mode is cost-effective option in both RB diagnosis and active lesion identification progress.

The tornado diagram shows the parameters that had the greatest impacts on base-case outcomes and how much they influenced the outcomes (**Supplementary Figure S5**). Probabilistic sensitivity analyses were conducted by taking 10000 random draws surrounding the basic values (**Supplementary Figure S6**). According to cost-effectiveness acceptability curves, at the current cost-effectiveness threshold, DLA-RB based diagnosis mode had a 100% probability of being the more cost-effective option in the process of RB diagnosis from all clinical-suspected retinoblastoma, and it had a 59% probability to be cost-effective for identification of active retinoblastoma from all treated retinoblastoma patients.

## Discussion

In the current study, we developed Deep Learning Assistant for Retinoblastoma (DLA-RB) that could accurately identify active retinoblastoma from the normal fundus and stable retinoblastoma fundus. The DLA-RB further exhibited superior sensitivity in identifying newly onset retinoblastoma and similar sensitivity in identifying relapse retinoblastoma. The diagnosis accuracy and sensitivity were superior/non-inferior than competent with about 2-5 years of experience in EUAs examination. Compared with referral procedures to ophthalmologic centers, DLR-RB based approach is cost-effective and readily implemented in the regional hospital.

According to the consensus from the American Association of Ophthalmic Oncologists and Pathologists, children with a family history of the retinoblastoma are highly recommended to take sufficient retina examination until the age of 7.^18^ Among all family members of retinoblastoma patients, offspring of bilateral have near 50% risk. In contrast, other siblings and nieces/nephews have less than 2.5% risk of harboring RB1 mutation. EUA is strongly recommended for any child unable to participate in an office examination sufficiently to allow a thorough examination of the retina.^18^ However, the frequent screening schedule is often limited by the scarcity of ophthalmological resources, especially in underdeveloped regions.^19^ Here, we proposed a novel screening solution for offspring and family members of patients. DLA-RB showed high efficiency with 0.991 of AUC and 0.979 sensitivity in the prospective validation dataset. The diagnostic accuracy and sensitivity of DLA-RB surpassed competent ophthalmologists. In real-world practice, the DLA-RB could provide automatic and real-time feedback on retinoblastoma screening, especially among the high-risk population.

Nowadays, although advanced multidisciplinary management concepts have been adopted in developed countries, less significant improvement in globe salvage rate was seen in the lower-middle-income country.^20^ DLA-RB could also be used as a screening technique for the contralateral eye of unilateral retinoblastoma patients who received primary enucleation. In a retrospective analysis comprising unilateral-presenting Group D retinoblastoma patients, during a median time of approximately 5 years follow-up, only 1 out of 55 developed a new tumor in the contralateral eye.^8^ Using DLA-RB at the local hospital could save time and financial burden when ophthalmologic centers are not near at hand.

Through multidisciplinary management, the globe salvage rate in the high-income country has tremendously improved from 34% in 1980–89 to 70% in 2010–20.^20^ Based on a meta-analysis, retinoblastoma recurrence occurs in about 15% of patients.^7^ Advanced retinoblastoma (IIRC Group D-E) has a significantly higher risk of recurrence.^21^ Extensive recurrence of subretinal or vitreous seeds was the most common cause of treatment failure and final enucleation. Thus, there are increasing burden of tumor surveillance after intraocular local control. Here, DLA-RB showed high sensitivity to detect active retinoblastoma in the background of stable retinoblastoma appearance (calcification, atrophy, laser and cryotherapy scar, and fibrosis). In real-world practice, DLA-RB can provide automatic assistance in treatment decision-making.

In the past two decades, telemedicine technology has enabled the centralization of care at a single center in developing countries to achieve patient outcomes comparable to those of developed countries.^22^ Within the domain of telemedicine, the DLA-RB can largely contribute to lowering the burden in diagnosis and follow-up and concentrate the limited healthcare resource in making multidisciplinary management. DLA-RB can also help to promote the availability of retinoblastoma screening and surveillance by improving the early diagnosis of population at-risk and long-term follow-up for retinoblastoma patients. Such an approach has successfully managed retinopathy of prematurity, a leading cause of childhood blindness worldwide. ^15,16,23^ Despite these remarkable results, some inherent limitations of DLA-RB are worth highlighting. First, the development and prospective validation of DLA-RB were conducted in a single institution (Beijing Tongren Hospital). The generalizability of diagnostic performance needs further validation. Yet, in the current study, fundus images in the present study were captured by five ophthalmological experts and two different commercially available cameras, which fully reflected the variability in operators’ experience, image quality, resolution, different camera systems, illumination, and field of view. Thus, the result from the perspective validation dataset can represent the performance of DLA-RB in real-world practice. Secondly, retinoblastoma begins as a round, translucent, gray to white tumor in the retina, similar to the halation caused by microbubble in couplant. Careful image quality control was required for ophthalmologists. An artificial intelligence-based quality control module is needed to incorporate with DLA-RB, which warrants further development. Third, accurately identifying active retinoblastoma also rely on a thorough examination of the posterior pole and peripheral retina, which depends on the experience of the operator. A deep-learning-based operation indicator may need to cooperate with DLA-RB to lower the risk of false-negative chance. Fourth, because of the low incidence of retinoblastoma, only 103 patients were included in prospective validation, which limited subgroup analysis. It warrants further study to explore the diagnosis accuracy of DLA-RB in different IIRC group retinoblastoma.

In conclusion, DLA-RB achieved high accuracy and sensitivity in identifying active retinoblastoma from the normal fundus and stable retinoblastoma fundus. Compared with referral procedures to ophthalmologic centers, DLA-RB based automatic screening and activity surveillance is also cost-effective. In the future, DLA-RB can incorporate telemedicine programs to reduce the burden in diagnosis and follow-up and concentrate the limited health-care resource in making multidisciplinary management.

## Methods

### Clinical trial registration and ethics approval

This single-center diagnostic study was done in Beijing Tongren hospitals in China. The methods were performed in accordance with relevant guidelines and regulations and approved by the Medical Ethics Committee of Beijing Tongren Hospital. Because individually identifiable information was removed during retrospective collection, written informed consent was exempted in the retrospective collected dataset. In the prospectively collected validation dataset, written informed consent was obtained from all caregivers prior to their inclusion. This study was registered on ClinicalTrials.gov (NCT05308043) on February 2022. Because this study was a diagnostic study that did not assign designative interventions to participants, clinical trial protocol is not applicable.

### Dataset collection

Training and internal test dataset were retrospectively collected between March 2018 and January 2022. All EUAs were performed for screening or pretreatment examination in daily clinical practice. Five ophthalmological experts (HS Zhao, X Ge, XL Xu, LB Jiang, JS, and JM Ma) obtained images using a commercially available camera (RetCam3; Natus Medical Incorporated) and the standard imaging protocol. In brief, posterior pole and 12 clocks of peripheral retina images were obtained during EUAs. Repeatedly images were taken if necessary. We collected all EUAs images from patients clinically diagnosed with retinoblastoma. For patients with several EUAs, we included all images in the retrospective dataset. Patients who were considered with Coats’ disease, persistent fetal vasculature and other retinal diseases were excluded.

The validation dataset was prospectively collected from February 2022 to June 2022. Clinically suspected retinoblastoma and treated retinoblastoma patients that first visited Beijing Tongren hospitals were standard EUAs. Patients involved in algorithm development and considered to have other retinal diseases were excluded. During prospective collection, a new generation of a commercially available camera (RetCam3) was deployed in Beijing Tongren hospital with updated illumination and imaging capture system. Thus, the prospective validation dataset was further divided into two subsets (Equipment 1 and 2) to examine the reliability of new equipment.

### Image quality control and labeling

All EUAs images were stored in jpeg format in the imaging databases. Poor-quality images resulting from halation, blur, and defocus, as well as non-EUAs images, were excluded manually. In addition, blurred images due to leukoplakia and secondary cataract in advanced retinoblastoma were also excluded. A multidisciplinary management team consisting of ophthalmology, pediatric and radiologist experts made personal treatment strategies after EUAs. The personal treatment strategies comprised a combination of a systemic chemotherapy regimen of carboplatin, vincristine, etoposide, and focal consolidation therapy. Each chemotherapy cycle was repeated every 3 to 4 weeks for 6 to 8 cycles, according to condition and tumor status. Follow-up by EUAs was undertaken before each cycle of chemotherapy and every 3 to 4 weeks thereafter, during which the adjuvant laser therapy and cryotherapy were applied as needed. In accordance with treatment strategies, five ophthalmological experts (HS Zhao, X Ge, XL Xu, LB Jiang, JS, and JM Ma) with a minimum of 15 years of experience labeled all images as “normal fundus”, “stable retinoblastoma” that specific treatment is not required, and “active retinoblastoma” that specific treatment is required.

### Development of DLA-RB

The patients from the retrospective dataset were randomly split into training and the internal validation dataset as five-fold cross-validation for developing and evaluating the performance of DLA-RB, respectively. To automatically distinguish “normal fundus”, “stable retinoblastoma”, and “active retinoblastoma”, we first compared the performance of some architectures including ResNet-50 and InceptionV3.^24-26^ ResNet-50 which did not occupy much computational resource achieved better performance and was chosen to complete the task. The input of the model was an image from EUAs. The output of the first model is a binary output determined whether the input image contained active retinoblastoma was among normal fundus and active retinoblastoma. The second binary classification task for determining among stable retinoblastoma and active retinoblastoma, whether the input image contained active retinoblastoma. In addition, the training dataset is imbalanced, so we adopted a class weight policy in the training process. All models were developed with Tensorflow 1.10.0 and Keras 2.2.4 on the server with three NVIDIA 1080 GPUs (Graphical Processing Units).^27^ All images were resized to 256×256 and then fed into models to train or test. The optimization algorithm was SGD (Stochastic Gradient Descent) ^28^, the default hyperparameters in Keras 2.2.4 were used, and at the same time, the batch size was 32. Besides, class weight was used for trading off the effect of the imbalanced distribution of two classes. Based on the repeated experiments, different epochs were also applied to train the models without underfitting. Moreover, the best models in terms of validation accuracy is saved as the final deployed model.

### Validation of DLA-RB

We first validated the performance of DLA-RB in identifying retinoblastoma activity in patients using an internal validation dataset and a prospective validation dataset from Beijing Tongren Hospital. For further performance evaluation, two varying degrees of expertise (competent and trainee), masked to the patients’ demographics and final multidisciplinary management strategy, were asked to classify every patient in the prospective dataset independently, and their results were compared with those of DLA-RB. The competent ophthalmologists attended doctors with about 2-5 years of experience in general ophthalmology and EUAs examination. The trainee was a resident who had finished EUAs training.

### Cost-utility analysis

Two Markov models were built using TreeAge Pro (TreeAge Software; Williamstown, MA, USA) to compare the cost-effectiveness between traditional ophthalmologic centers-based diagnosis mode and DLA-RB based diagnosis mode from a societal perspective. For the first binary model, we simulated a hypothetical cohort of a newborn with RB through 5 1-year Markov cycles. With the growth of age, children could be found to have eye symptoms by their parents according to specific probabilities and then go to the hospital. Diagnosed patients received routine treatment and management, whereas undiagnosed patients could still be examined in the next cycle. According to the Management Guidelines for Childhood Screening for Retinoblastoma Families, we determined the screening intervals for undiagnosed patients.^18^ For the second binary model, simulated a hypothetical cohort of 2-year-old RB patients who have received regular treatment and were in an inactive stage through a total of 3 1-year Markov cycles. They were routinely reviewed every three months, and those who had active lesions received further treatment.

Parameters used in Markov models were collected from our study and previous studies. Examination and treatment costs were collected in Chinese yuan from Beijing Tongren Hospital and converted to US dollars at an exchange rate of 6.45 yuan per dollar (**Supplementary Table S4**). Both direct and indirect costs were included, and the specific composition of costs and annual costs of traditional ophthalmologic centers-based diagnosis mode and DLA-RB-based diagnosis mode was shown in **Supplementary Table S5-6**. Under traditional ophthalmologic centers-based diagnosis mode, the patient and one accompanying family member spent more time in tertiary hospitals or eye hospitals, resulting in transportation, accommodation, and food costs. In DLA-RB-based diagnosis mode, we charged an additional $15.5 for each use. Only one accompanying family member’s wage loss was counted.

Primary results were incremental cost-utility ratios (ICURs), which were calculated using the following formula:

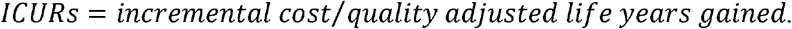

China’s per capita gross domestic product (GDP) in 2021 was $12551. According to the WHO definition, intervention costing between 1 to 3 times the per capita GDP was cost-effective, costing less than the per capita GDP was highly cost-effective, and costing more than three times the per capita GDP was not cost-effective. ^24^ Both 1-way deterministic and simulated probabilistic sensitivity analyses were performed to test the sensitivity and robustness of base-case values, the uncertainty ranges of parameters were presented in **Supplementary Table S7**.

### Visualization and statistical analysis

To visualize the decision ways of the model used, we applied the Grad-CAM to generate heatmaps.^25^ The performance of DLA-RB was estimated by accuracy, sensitivity, specificity, positive predictive value, negative predictive value, and F1 score for the identification of each category. We used the receiver operating characteristic (ROC) curve and precision-recall curve to show the diagnostic performance of the DLA-RB in discriminating binary classification tasks. N-out-of-N Bootstrapping with 1000 replicates was used to estimate 95% confidence intervals (95% CI) of the performance metrics at eye level. Human ophthalmologists’ performance was compared with 95% CI of DLA-RB.

All statistical analysis was performed using R Statistical Software (version 4.1.1; R Foundation for Statistical Computing, Vienna, Austria), Stata (17.0, StataCorp LLC, College Station, TX).

### Data availability

Python scripts enabling the main steps of the analysis are available from the corresponding author on reasonable request. The data and materials in this study are available from the corresponding author on reasonable request.

## Data Availability

All data produced in the present study are available upon reasonable request to the authors

## Acknowledgments

The authors thanks Beijing Hospitals Authority’ Ascent Plan (DFL20190201); National Natural Science Foundation of China (82141128); The Capital Health Research and Development of Special (2020-1-2052); Science & Technology Project of Beijing Municipal Science & Technology Commissi on (Z201100005520045, Z181100001818003) for supporting this study.

## Contributors

JMM, RHZ and DL contributed to the concept of the study. JMM, WBW critically reviewed the manuscript. RHZ, LD, RYL,KZ, and YTL designed the study and did the literature search. RHZ, LD, RYL,KZ,YTL, HSZ, JTS, XG, XLX, LBJ, XHS, CZ, WDZ, LYX, HTW, HYL, CYY, and JL collected the data. RHZ, LD, RYL,KZ, andYTL contributed to the design of the statistical analysis plan. RHZ, LD, and KZ did the data analysis and data interpretation. RHZ and RYL drafted the manuscript. JMM and WBW provided research funding, coordinated the research, and oversaw the project. All authors had access to all the raw datasets and the corresponding authors (JMM and WBW) has verified the data and had final decision to submit for publication. All authors reviewed and approved the final manuscript

