## Supplementary Tables and figures for "Automatic Retinoblastoma Screening and Surveillance Using Deep Learning": Appendix.docx

**Figure S1.** Typical low-quality images that manually excluded.

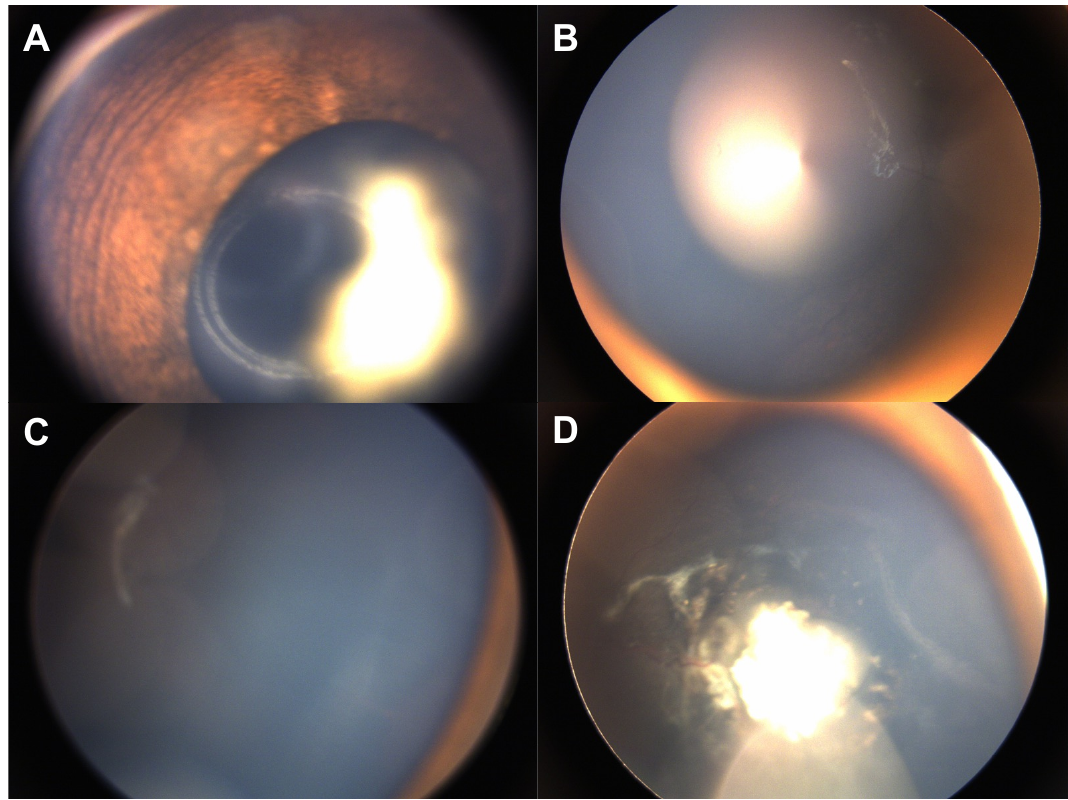

Low-quality images due to non-fundus image (A), halation (B), blurs (C), and defocus (D).

**Figure S2.** Receiver operating characteristic curves compared ResNet-50 and InceptionV3

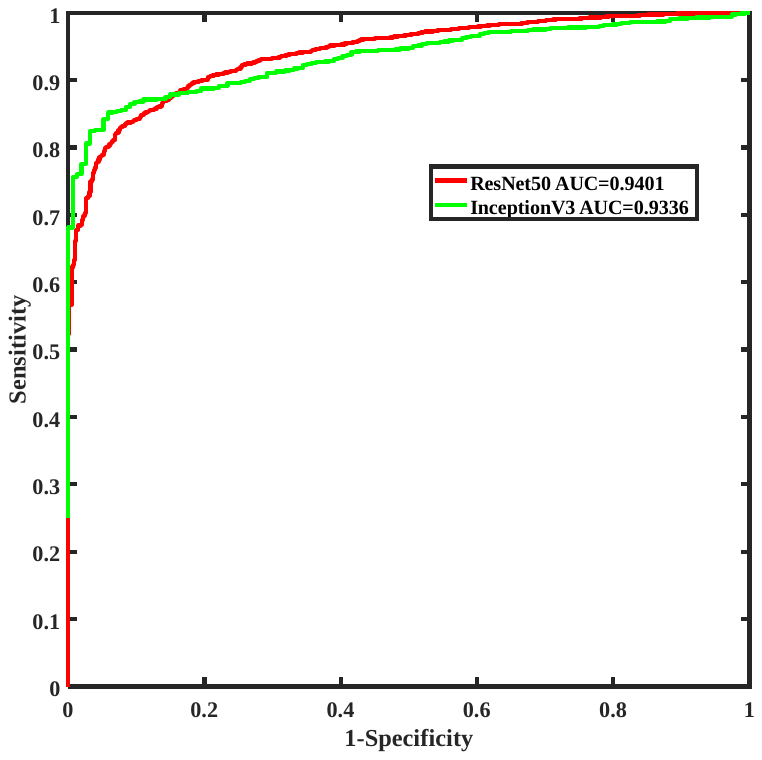

ResNet-50 and InceptionV3 were trained to distinguish stable and active RB lesion on fundus image. In five fold cross-validation, receiver operating characteristic curves were used to compared performance of these two architectures.

**Figure S3.** Receiver operating characteristic curves of DLA-RB in distinguishing normal fundus and active lesion.

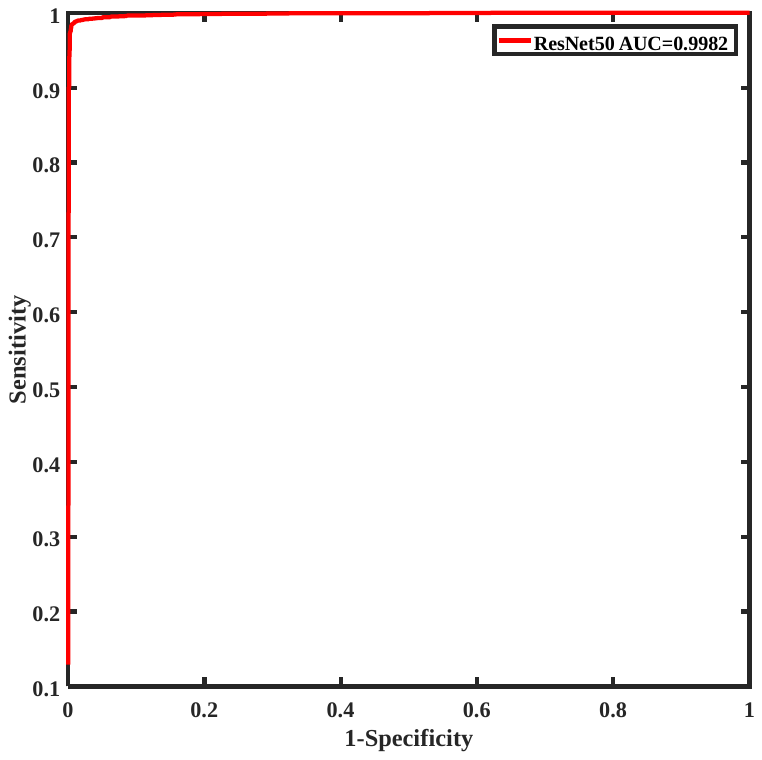

^1^

**Figure S4.** In normal-active retinoblastoma identification, the fundus images of misclassification by DLA-RB.

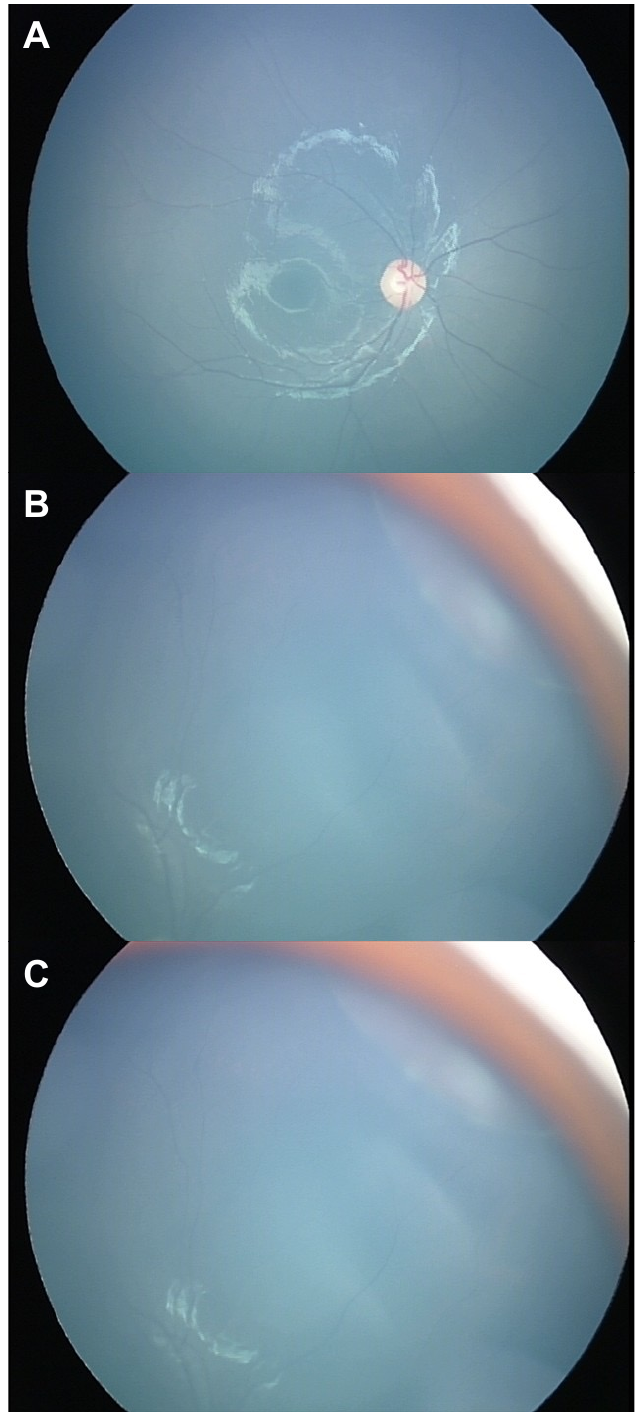

A, posterior fundus. B-C, the retinoblastoma tumor located at superior-nasal

**Figure S5.** Results of deterministic 1-way sensitivity analysis.

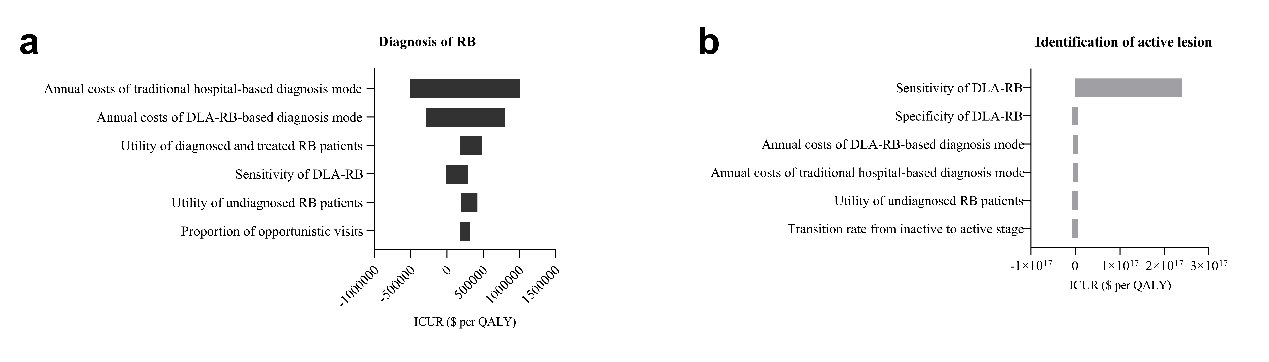

DLA-RB, deep Learning Assistant for Retinoblastoma Monitoring; ICUR, incremental cost-utility ratio; QALY, quality-adjusted life-year.

**Figure S6.** Probabilistic sensitivity analysis.

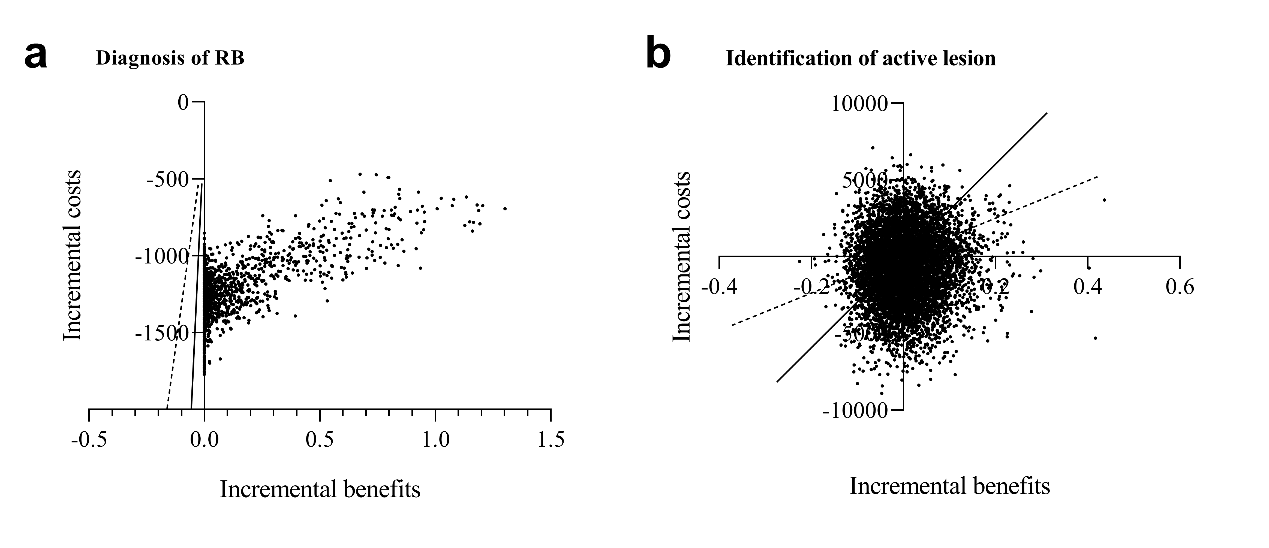

Costs are given in US dollars. Incremental benefits are defined as incremental QALYs. Dashed and solid lines represent one-time and three-times GDP, respectively. QALY= quality-adjusted life-year; GDP= gross domestic product; RB, retinoblastoma.

**Figure S7.** Cost-effectiveness acceptability curve.

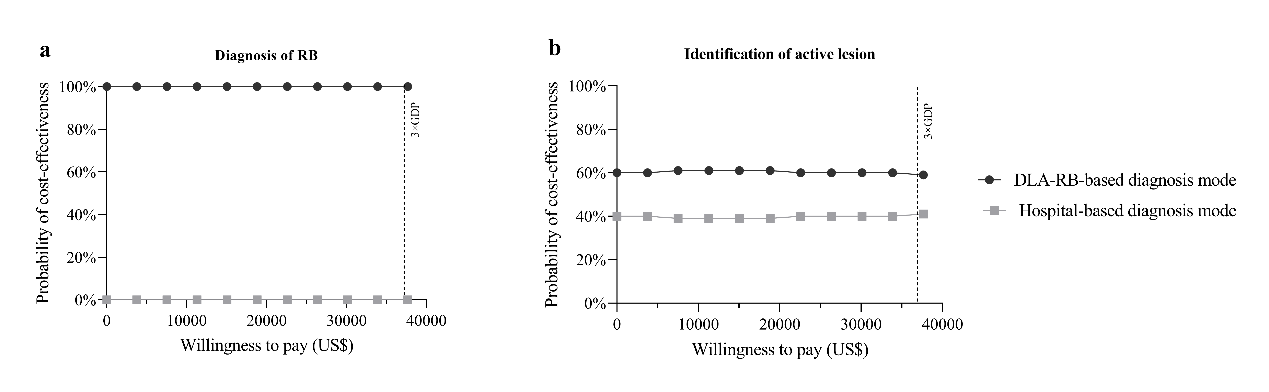

RB, retinoblastoma; GDP= gross domestic product.

**Table S1**. Basic characteristics of patients in development dataset.

|  | Development dataset |
| --- | --- |
|  | n=713 |
| Age at diagnosis |  |
| <12month | 97 (13·4%) |
| 12-23 month | 200 (28·1%) |
| 24-35 month | 175 (24·6%) |
| ≥36 month | 240 (33·7%) |
| Average number of exams median (interquartile range) | 2 (1-4) |

**Table S2**. Performance of human reader in prospective validation dataset (Normal-active lesion identification task).

|  | Sensitivity (95% CI) | Specificity (95% CI) | Positive predictive value (95% CI) | Negative predictive value (95% CI) |
| --- | --- | --- | --- | --- |
| Reader 1 | 0·848 [0·711-0·973] | 0·986 [0·922-1·000] | 0·975 [0·868-0·999] | 0·907 [0·817-0·962] |
| Reader 2 | 0·522 [0·369-0·671] | 1·000 [0·946-1·000] | 1·000 [0·858-1·000] | 0·753 [0·650-0·838] |
| Reader 3 | 0·522 [0·369-0·671] | 1·000 [0·946-1·000] | 1·000 [0·858-1·000] | 0·758 [0·657-0·842] |
| Reader 4 | 0·804 [0·661-0·906] | 0·754 [0·635-0·849] | 0·685 [0·544-0·805] | 0·852 [0·738-0·930] |
| Reader 5 | 0·543 [0·390-0·691] | 1·000 [0·948-1·000] | 1·000 [0·863-1·000] | 0·767 [0·666-0·849] |
| Reader 6 | 0·913 [0·792-0·976] | 1·000 [0·948-1·000] | 1·000 [0·916-1·000] | 0·945 [0·866-0·985] |

Competent ophthalmologists: Reader 1, 4 and 6; trainee ophthalmologists: Reader 2, 3 and 5.

**Table S3**. Performance of human reader in prospective validation dataset (Normal-active lesion identification task).

|  | Sensitivity (95% CI) | Specificity (95% CI) | Positive predictive value (95% CI) | Negative predictive value (95% CI) |
| --- | --- | --- | --- | --- |
| Reader 1 | 0·848 [0·711-0·973] | 0·938 [0·711-0·837] | 0·975 [0·868-0·999] | 0·682 [0·451-0·861] |
| Reader 2 | 0·522 [0·369-0·671] | 0·938 [0·698-0·998] | 0·960 [0·796-0·999] | 0·405 [0·248-0·579] |
| Reader 3 | 0·522 [0·369-0·671] | 0·688 [0·413-0·890] | 0·828 [0·642-0·942] | 0·333 [0·180-0·942] |
| Reader 4 | 0·804 [0·661-0·906] | 0·804 [0·661-0·906] | 0·822 [0·679-0·920] | 0·471 [0·230-0·722] |
| Reader 5 | 0·543 [0·390-0·691] | 1·000 [0·794-1·000] | 1·000 [0·863-1·000] | 0·432 [0·271-0·605] |
| Reader 6 | 0·913 [0·792-0·976] | 0·975 [0·617-0·984] | 0·955 [0·845-0·994] | 0·778 [0·524-0·936] |

Competent ophthalmologists: Reader 1, 4 and 6; trainee ophthalmologists: Reader 2, 3 and 5.

**Table S4.** Parameters used in Markov models.

|  | **Diagnosis of RB** | **Identification of active lesion in treated retinoblastoma patients** |
| --- | --- | --- |
| Proportion of opportunistic visits among…^2^ |  |  |
| 0–1-year-old patients | 31% | NA |
| 1-2-year-old patients | 37% | NA |
| 2-3-year-old patients | 20% | NA |
| 3-4-year-old patients | 9% | NA |
| 4-5-year-old patients | 3% | NA |
| Transition rate from inactive to active stage every three months^3,4^ | NA | 0·53% |
| Sensitivity of DLA-RB | 97·92% | 97·83% |
| Specificity of DLA-RB | NA | 80% |
| Utility of… |  |  |
| Undiagnosed RB patients^5,6^ | 0·5514 | 0·5514 |
| Diagnosed and treated RB patients^5,6^ | 0·701 | 0·701 |
| Annual costs of… ($) |  |  |
| Traditional ophthalmologic centers-based diagnosis mode |  |  |
| 0–1-year-old patients | 10944 | NA |
| 1-2-year-old patients | 4690 | NA |
| 2-3-year-old patients | 3127 | 782 |
| 3-4-year-old patients | 2345 | 782 |
| 4-5-year-old patients | 1563 | 782 |
| DLA-RB-based diagnosis mode |  |  |
| 0–1-year-old patients | 7859 | NA |
| 1-2-year-old patients | 3368 | NA |
| 2-3-year-old patients | 2246 | 561 |
| 3-4-year-old patients | 1684 | 561 |
| 4-5-year-old patients | 1123 | 561 |
| Discount rate^4^ | 3·5% | 3·5% |

DLA-RB, deep Learning Assistant for Retinoblastoma Monitoring; DLA, Deep Learning Assistant; NA, not applicable.

Costs are given in US dollars

**Table S5.** Cost computation of once retinoblastoma examination using both modes·

|  | **Traditional ophthalmologic centers-based diagnosis mode ($)** | **DLA-RB-based diagnosis mode ($)** |
| --- | --- | --- |
| Direct medical costs |  |  |
| Outpatient preoperative examination fees | 155·04 | 155·04 |
| Hospitalization fees | 310·08 | 310·08 |
| Expenses for the use of DLA-RB-based diagnosis mode | 0 | 15·5 |
| Direct non-medical costs |  |  |
| Accommodation | 46·51 | 0 |
| Foods | 46·51 | 0 |
| Daily necessities and Transportation | 31 | 31 |
| Indirect costs | 99·52 | 49·76 |
| Societal costs per person ($) | 781·68 | 561·38 |

DLA-RB, deep Learning Assistant for Retinoblastoma Monitoring; NA, not applicable.

**Table S6.** Annual Costs retinoblastoma examination using both modes.

|  | **Traditional ophthalmologic centers-based diagnosis mode ($)** | **DLA-RB based diagnosis mode ($)** |
| --- | --- | --- |
| 0–1-year-old patients | 10944 | 7859 |
| 1-2-year-old patients | 4690 | 3368 |
| 2-3-year-old patients | 3127 | 2246 |
| 3-4-year-old patients | 2345 | 1684 |
| 4-5-year-old patients | 1563 | 1123 |

DLA-RB, deep Learning Assistant for Retinoblastoma Monitoring

**Table S7.** Base-case values and uncertainty ranges of parameters in Markov models.

|  | **Diagnosis of RB** | | | **Identification of active lesion in treated retinoblastoma patients** | | |
| --- | --- | --- | --- | --- | --- | --- |
|  | Base-case value | Range for one-way sensitivity analysis | Distributions used in the probabilistic sensitivity analysis | Base-case value | Range for one-way sensitivity analysis | Distributions used in the probabilistic sensitivity analysis |
| Proportion of opportunistic visits among… | | | | | | |
| 0–1-year-old patients | 31% | ±10% (28%, 34%) | Beta (68·69,152·89) | NA |  |  |
| 1-2-year-old patients | 37% | ±10% (33%, 41%) | Beta (62·63,106·64) | NA |  |  |
| 2-3-year-old patients | 20% | ±10% (18%, 22%) | Beta (79·8,319·2) | NA |  |  |
| 3-4-year-old patients | 9% | ±10% (81%, 99%) | Beta (9·1,1·01) | NA |  |  |
| 4-5-year-old patients | 3% | ±10% (27%, 33%) | Beta (69·7,162·63) | NA |  |  |
| Transition rate from inactive to active stage every three months | NA |  |  | 0·53% | ±10% (0·48%, 0·58%) | Beta (99·46,18667·46) |
| Sensitivity of DLA-RB | 97·92% | (92·7%, 100%) | Beta (1·1,0·02) | 97·83% | (93·2%, 100%) | Beta (1·19,0·03) |
| Specificity of DLA-RB | NA |  |  | 80% | (55·6%, 100%) | Beta (19·2,4·8) |
| Utility of… | | | | | | |
| Undiagnosed RB patients | 0·5514 | ±10% (0·5, 0·61) | Beta (44·31,36·05) | 0·5514 | ±10% (0·5, 0·61) | Beta (44·31,36·05) |
| Diagnosed and treated RB patients | 0·701 | ±10% (0·63, 0·77) | Beta (29·2,12·45) | 0·701 | ±10% (0·63, 0·77) | Beta (29·2,12·45) |
| Annual costs of… ($) | | | | | | |
| Traditional ophthalmologic centers-based diagnosis mode | | | | | | |
| 0–1-year-old patients | 10944 | ±20% (8755, 13133) | Gamma (16,684) | NA |  |  |
| 1-2-year-old patients | 4690 | ±20% (3752, 5628) | Gamma (16,293·13) | NA |  |  |
| 2-3-year-old patients | 3127 | ±20% (2502, 3752) | Gamma (16,195·44) | 782 | ±20% (626, 938) | Gamma (16,48·88) |
| 3-4-year-old patients | 2345 | ±20% (1876, 2814) | Gamma (16,146·56) | 782 | ±20% (626, 938) | Gamma (16,48·88) |
| 4-5-year-old patients | 1563 | ±20% (1250, 1876) | Gamma (16,97·69) | 782 | ±20% (626, 938) | Gamma (16,48·88) |
| DLA-RB-based diagnosis mode | | | | | | |
| 0–1-year-old patients | 7859 | ±20% (6287, 9431) | Gamma (16,491·19) | NA |  |  |
| 1-2-year-old patients | 3368 | ±20% (2694, 4042) | Gamma (16,210·5) | NA |  |  |
| 2-3-year-old patients | 2246 | ±20% (1797, 2695) | Gamma (16,140·38) | 561 | ±20% (449, 673) | Gamma (16,35·06) |
| 3-4-year-old patients | 1684 | ±20% (1347, 2021) | Gamma (16,105·25) | 561 | ±20% (449, 673) | Gamma (16,35·06) |
| 4-5-year-old patients | 1123 | ±20% (898, 1348) | Gamma (16,70·19) | 561 | ±20% (449, 673) | Gamma (16,35·06) |
| Discount rate | 3·5% | NA | NA | 3·5% | NA | NA |

RB, retinoblastoma; NA, not applicable.
